# Protocol for longitudinal assessment of SARS-CoV-2-specific immune responses in healthcare professionals in Hannover, Germany: the prospective, longitudinal, observational COVID-19 Contact (CoCo) study

**DOI:** 10.1101/2020.12.02.20242479

**Authors:** Alexandra Jablonka, Christine Happle, Anne Cossmann, Metodi V. Stankov, Anna Zychlinsky Scharff, Diana Ernst, Georg M.N. Behrens

## Abstract

**Background:** During the current pandemic, healthcare professionals (HCP) have been at the frontline of the crisis. Serological screening may help in identifying severe acute respiratory syndrome coronavirus 2 (SARS-CoV-2) prevalence. However, given the rapidly evolving situation in spring 2020, many questions regarding coronavirus disease 2019 (COVID-19) infection risk and utility of serological testing remained unanswered. To address these questions, we initiated the COVID-19 Contact (CoCo) study at Hannover Medical School, a large university hospital in Northern Germany and affiliated care providers.

**Methods:** The CoCo study is an ongoing, prospective, longitudinal, observational study in HCP and individuals with potential contact to SARS-CoV-2. It monitors anti-SARS-CoV-2 immunoglobulin serum levels and collects information on symptoms of respiratory infection, work and home environment, and self-perceived SARS-CoV-2 infection risk. Inclusion criteria are (1) working as HCP in clinical care at our university centre, affiliated hospitals or private practices, (2) written informed consent and (3) age >18 years. Exclusion criteria are (1) refusal to give informed consent and (2) contraindication to venepuncture. Study participants are asked to provide weekly to six-monthly samples (7.5 ml serum and 7.5 ml EDTA blood) and fill out a questionnaire. Since March 2020, around 1250 HCP have been included in the study. At each study visit, sera are screened for anti-SARS-CoV-2 spike protein 1 (S1) immunoglobulin G (IgG) by enzyme-linked immunosorbent assay (ELISA). Positive or borderline positive samples are re-assessed with an alternative serological test. Individual results for each study participant are made available online via a dedicated study website. This study also aims to compare different serological testing assays, as well as explore further humoral and cellular immune markers. Study protocols are continually adapted to the rapidly evolving situation of the current pandemic.

**Discussion:** This ongoing prospective study will aim to answer central questions on the prevalence and kinetics of anti-SARS-CoV-2-humoral immune responses and the validity of serological testing of HCP in a region with high healthcare standard and comparatively low COVID-19 prevalence. As such, our results are highly relevant to other regions and may support HCP around the world in managing this unprecedented situation.

**Trial registration:** German Clinical Trial Registry, DRKS00021152. Registered 4th April 2020 -retrospectively registered, https://www.drks.de/drks_web/navigate.do?navigationId=trial.HTML&TRIAL_ID=DRKS00021152

**Protocol summary:** 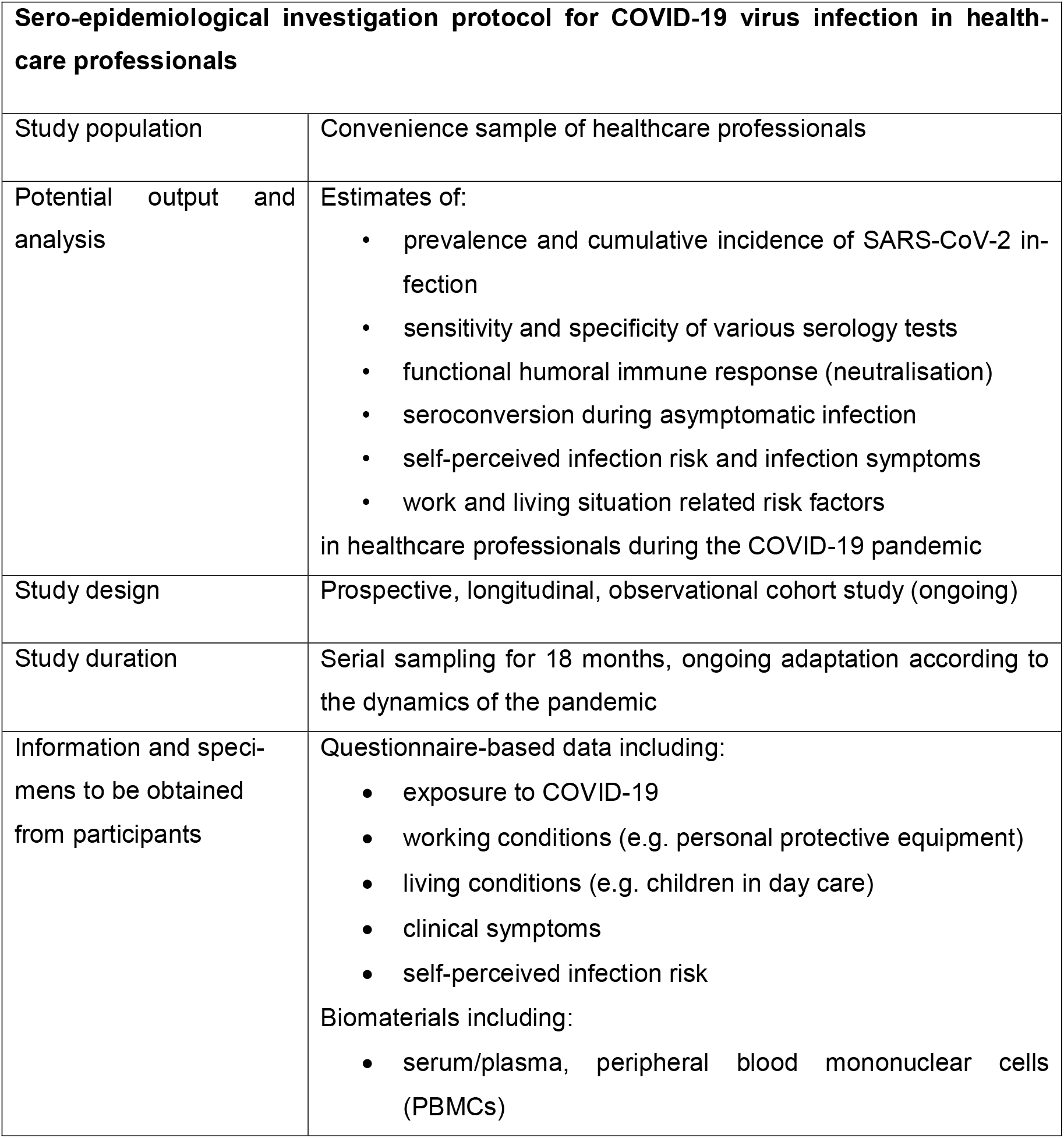

## Background

In late 2019, the severe acute respiratory syndrome coronavirus 2 (SARS-CoV-2) emerged in Wuhan, China, and has subsequently spread throughout the world [1]. Since the beginning of the current pandemic, the risk to healthcare professionals (HCP) of contracting this infection while taking care of patients has been extensively discussed [2, 3]. During the early phase of the pandemic, when personal protective equipment (PPE) and other protective measures were scarce, considerable numbers of HCP in high incidence regions such as Hubei, China or Lombardy, Italy were infected [3, 4]. During later phases, when many healthcare systems were better prepared, infection rates amongst HCP fell. Cases in healthcare workers now likely reflect pandemic spread in the general population [5, 6]. In Germany, the first confirmed case of coronavirus disease 2019 (COVID-19) was registered on January 6^th^, 2020 in Munich [7]. The return of German tourists from holiday in Austria and Northern Italy later spread the virus to 13 of 16 federal states within one month [8]. Initially there was an exponential increase in newly confirmed cases which flattened in April. Germany has reached a total of 204,964 positively tested cases on July 25th 2020 (mortality rate 4.5% [9]). As in many countries, this had significant impact on the German healthcare system, where healthcare provision shifted from routine clinical care to preparing for a potentially high number of severely ill COVID-19 cases [10-12]. By implementing social distancing measures, testing and contact tracing, the spread of infection was slowed before the healthcare system became overwhelmed.

Besides innate and T cell responses, B cells are central in the antiviral defence against coronavirus infections [13-15]. SARS-CoV-2 specific antibody levels correlate with disease severity and are usually detectable during the first three weeks after symptom onset [16-18]. Serological screening may help determine SARS-CoV-2 prevalence amongst HCP and possibly identify those with some level of immunity. However, given the rapidly evolving situation, many questions regarding the infection risk in HCP and the characterisation of COVID-19 immunity by serological analysis or other screening tools remain unanswered.

The main objectives of the here described study are to systematically determine the longitudinal prevalence and cumulative incidence of true SARS-CoV-2 infections and the self-perceived risk of contracting this virus in HCP. Also, we aim at identifying the rate of asymptomatic SARS-CoV-2 infections in HCP, to assess the sensitivity and specificity of different anti-SARS-CoV-2 serology test systems, and to answer further questions on the immune response against SARS-CoV-2 in HCP.

## Methods/Design

### Aims

In March 2020, we initiated the CoCo study at Hannover Medical School, a large university hospital in Northern Germany, to address central questions regarding the risk of COVID-19 in HCP and the utility of serological screenings for SARS-CoV-2 in HCP. The central aims leading to the initiation of this study are listed here:

Primary Objectives

- to systematically screen for anti-SARS-CoV-2 immunoglobulins in HCP, to determine the prevalence and cumulative incidence of SARS-CoV-2 infection
- to assess the self-perceived risk of having had SARS-CoV-2 infection and to identify potential risk factors in HCP

Secondary Objectives

- to identify asymptomatic SARS-CoV-2 infections in HCP by serology testing
- to determine the sensitivity and specificity of different anti-SARS-CoV-2 serology test systems
- to describe the kinetics, magnitude, and quality of anti-SARS-CoV-2 humoral and cellular immune response in HCP
- to develop a serology testing strategy for improving serology test result interpretation
- to compare the anti-SARS-CoV-2 prevalence in the CoCo study with regional dynamic of COVID-19 cases.

### Setting

Hannover Medical School is a university hospital located in the capital of Lower Saxony, Germany with 1,520 beds and 7,500 employees, of which 3,100 work as HCP. In 2019, our hospital reported 62,000 inpatient and 525,000 outpatient cases. Hannover Medical School has clinical and research focuses on immunology and transplantation, regeneration, pneumology, and infectious diseases and is part of the nationally funded clinical research initiatives German Center for Infection Research and German Center for Lung Research.

The CoCo study was initiated in March 2020, shortly before the local peak of the pandemic, when the first COVID-19 related restrictions in Germany were imposed and social distancing and other measures to mitigate viral spread were enforced nationwide (Fig. 1). During this time, a massive reduction in routine as well as emergency medical care occurred [19]. During the national ‘lockdown’, PPE and infection control protocols at Hannover Medical School were ramped up hospital-wide. An interdisciplinary ‘corona task force’ was set up to coordinate clinical approaches regarding COVID-19 and to inform hospital staff via regular emails on the number of treated COVID-19 patients in our hospital and all measures and policies related to the pandemic. Comparably few COVID-19 cases occurred in our region and were treated in our hospital, as illustrated in Figure 1. No systematic testing of hospital staff was initiated, and employees were asked to seek medical attention and get tested for SARS-CoV-2 according to current national guidelines. Starting from the end of March 2020, hospital staff can undergo on-site SARS-CoV-2 nasopharyngeal swab testing when an infection is suspected.

**Fig. 1.**
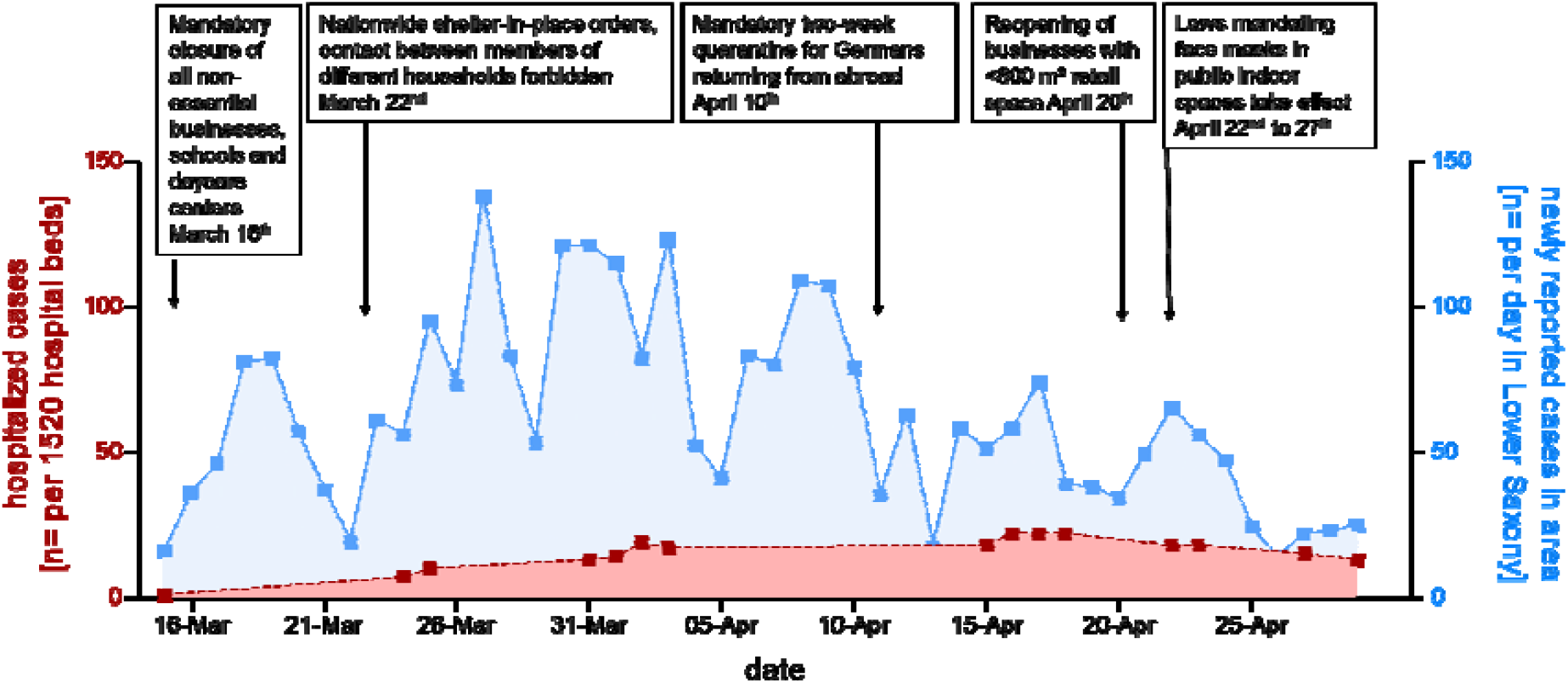
Local infection and inpatient COVID-19 patients during COVID-19 related lockdown in Germany: Newly reported infections with SARS-CoV-2 in the region of Hannover (blue) and total number of COVID-19 patients treated at Hannover Medical School (red) including dates of COVID-19 related lockdown strategies in Germany.

### Study population

The study includes frontline HCP involved at all levels of patient care. Participants from departments both with high COVID-19 exposure (e.g. intensive care units, emergency department, infectious disease unit, respiratory medicine) and other departments (i.e. psychosomatic department) of Hannover Medical School are recruited. Every attempt is made to include a representative sample in terms of profession, exposure and age of personnel. The study also includes HCP in hospitals affiliated with Hannover Medical School or in private practices. Furthermore, a positive control cohort consisting of patients with polymerase chain reaction (PCR)-confirmed COVID-19 and their relatives or individuals with close contact to these patients is being recruited.

Eligibility criteria are as follows. Inclusion criteria are (1) working as HCP with direct patient contact in selected departments of Hannover Medical School, affiliated hospitals and private practices, (2) written informed consent, (3) age >18 years. Exclusion criteria are (1) refusal to give informed consent, (2) contraindication to venepuncture. Inclusion criteria for the positive control cohort are (1) previously or currently positive SARS-CoV-2 PCR from nasopharyngeal swab or direct contact with such a patient, (2) written informed consent, (3) age >18 years or written informed consent of a legal guardian. Exclusion criteria for the positive control cohort are the same as for the entire study population.

### Study design

The CoCo study is an ongoing, prospective cohort analysis which longitudinally monitors SARS-CoV-2 specific immunoglobulin G (IgG) serum levels and collects information on symptoms of respiratory infection, work and living environment, and self-perceived infection risk in HCP.

The pilot ‘CoCo cohort 1.0’ enrolled n=217 HCP from Hannover Medical School units involved in COVID-19 patient care from March 23^th^ until April 17^th^ 2020 [17]. Participants were informed about the opportunity to participate by a designated study team for each participating department, word of mouth, and the institution-wide COVID-19 email bulletin. No incentives for participation in the CoCo study were offered, but participants were given access to their anti-SARS-CoV-2 enzyme-linked immunosorbent assay (ELISA) results via web-based personalized access codes. After written informed consent was obtained, participants filled out a baseline questionnaire and blood samples were drawn by members of the research team or study participants themselves (7.5 ml serum and 7.5 ml EDTA tubes). After enrolment, participants of the CoCo 1.0 cohort were asked to provide serum specimens weekly during the first two months, followed by monthly testing. EDTA blood was collected at baseline and week 4 and for participants with positive ELISA results at the next study visit.

Enrolment for the ‘CoCo 2.0’ cohort is ongoing and began in May 2020 with recruitment of n>1,140 HCP thus far, from a wide variety of clinical departments. In this study, serological testing will be conducted every six months until June 2021. The frequency of sampling and duration of the study may be adjusted depending on the regional dynamic of COVID-19 prevalence and incidence of seroconversion in the ongoing ‘CoCo 1.0’ sampling. Figure 2 illustrates study arms as well as visit and sampling frequencies. Local COVID-19 prevalence and the frequency of positive ELISA results as well as interim data from questionnaires will be analysed regularly in order to optimize the scheduling of study visits and to minimize blood sampling.

**Fig. 2.**
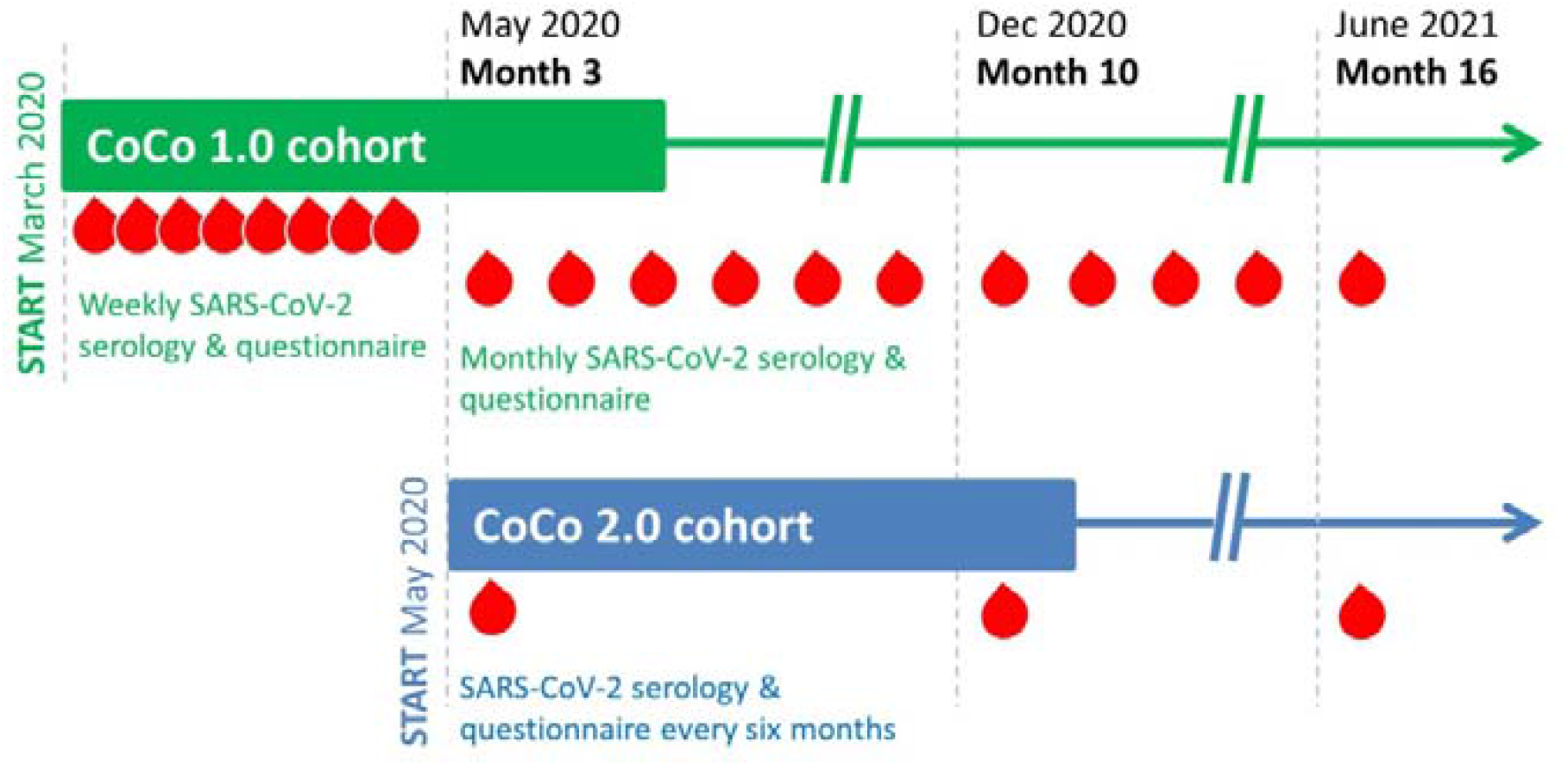
Study design and sampling frequency in the CoCo study.

### Biosampling and measures to prevent study visit-associated infections

In order to minimize the risk of transmission while conducting the study and in accordance with COVID-19 contact restrictions, several decentralized sites for blood sampling are established on the campus. Study participants are asked to adhere to social distancing and appropriate measures for infection prevention (e.g. face masks) when collecting or delivering study material. Enrolment and follow-up visits are primarily organized by designated members of participating departments without physical contact. Communication regarding study logistics is conducted primarily by telephone or email. All study personnel is trained in infection prevention and control measures (standard precautions as determined by national guidelines of the Robert Koch Institute, the German institute of public health, and local guidelines of the university hospital). These procedures include proper hand hygiene and use of surgical masks at all times and are universally implemented in our hospital to minimize the risk of spread among HCP and patients. All blood tubes and questionnaires as well as pseudonym codes are distributed to participants directly or through a designated member of each department coordinating study visits in that team. In addition to the provision of study materials at study sites on campus, study kits can also be shipped directly to participants. These kits are provided in depersonalized, relabelled boxes containing sampling material, questionnaires, and written informed consent forms. These kits are designed for use by participants that are in quarantine, by external study participants, e.g., from private practices, or by those absent from Hannover Medical School campus for other reasons. By using an on-demand prepaid express service, compliant with national standards for shipment of biological samples, these boxes can be mailed from anywhere in Germany at any time without disclosure of the sending party’s address, which ensures adherence to data safety regulations and quick delivery of study materials to the laboratory. Sample collection is conducted from Monday to Wednesday in the week of study visits, and anti-SARS-CoV-2 S1 IgG ELISAs are performed regularly on Thursday of that study week. All personnel involved in collection and transport of specimens is trained in safe handling practices. For each biological sample, time of collection, conditions of transport and time of arrival at the study laboratory are recorded. Test results are provided online on Friday of the study week through a personalised login. The CoCo study website is used to communicate test results and to inform study participants about news and developments concerning the study. Participants are able to contact the study team via email or telephone at any time to discuss questions regarding their results or participation.

### Data collection

The variables collected at the study visits are listed in Table 1. At baseline, HCP are asked to provide information about their age, sex and health (e.g. cardiovascular or respiratory diseases, regular medication etc.). At each study visit, questions regarding personal working and living conditions, type and area of work, number of persons and children living in the household, exposure to persons with suspected or confirmed SARS-CoV-2 infection etc. are asked. To assess the self-perceived probability of having contracted SARS-CoV-2, the following question is asked at each visit: ‘How high do you rate the likelihood of having been infected with SARS-CoV-2 so far? (0-100%)’. Furthermore, information on symptoms such as cough, fever, dyspnoea, rhinitis, sore throat, dysgeusia, and anosmia is collected. For each symptom, subjects are asked to provide a subjective scoring between 0 (absence of symptom) and 10 (highest imaginable intensity). The questionnaire will be adapted regularly to include novel and relevant information on COVID-19 related symptoms and risk factors.

**Table 1.** Participant information and questionnaire-based data collected in the CoCo study.

| Type of data | Collected variables |
| --- | --- |
| **General information** | Patient and place of work, date of enrolment, demographic data (e.g. age, sex, job title) |
| **Risk perception** | Self-perceived probability of having contracted a SARS-CoV-2 infection (rate 0-100%) |
| **Clinical history** | Chronic diseases, cardiovascular or respiratory illness, regular medication, immunosuppression, previous infections |
| **Complaints** | Cough, fever, dyspnoea, rhinitis, sore throat, dysgeusia and anosmia, other |
| **Work situation** | Department, area of work and position, indirect/direct contact with suspected or confirmed cases of COVID-19, sick leave quarantine (and endurance) |
| **Living situation** | Housing situation, children at day care, number and ages of all members of household |
| **Other data** | Travel history (recent return from high risk regions), contact with known COVID-19 patients, anxiety associated with risk of SARS-CoV-2 infection at work or outside of work. Level of personal burden due to the pandemic, level of adherence to social distancing rules, perception of disruption of personal routines through pandemic control measures |

### Laboratory setup and immunological analyses

The laboratory primarily analysing study samples is a research lab in the department for rheumatology and immunology at Hannover Medical School. Specimens are either collected from the on-campus study sites or express-mailed directly to the lab. Every biospecimen is processed, aliquoted and stored on the day of arrival. Primary serological testing is performed in the study lab. As mentioned above, Hannover Medical School is part of regional and national networks such as the German Centers for Infection and Lung Research, and further sample analyses will be performed in collaborating laboratories.

Besides regular testing for SARS-COV-2 IgG, which is performed in every participant in the week of each study visit, further analyses are planned for selected participants and time points (Table 2).

**Table 2.** Planned analyses of the CoCo study.

| Type of specimen | Planned analyses |
| --- | --- |
| **Serum/plasma** | ELISA for SARS-COV-2 S1 specific IgG/IgA<br>ELISA for SARS-COV-2 NCP specific IgG<br>SARS-CoV-2 rapid antibody test<br>*In vitro* cell culture-based virus neutralisation assay<br>Surrogate virus neutralisation test (sVNT)<br>[Analysis of inflammatory mediators, soluble angiotensin converting enzyme 2 receptor...] |
| **PBMCs** | Analysis of cellular immune responses against SARS-CoV-2 |

### Serological Testing

As a primary screening system, a semiquantitative ELISA for SARS-COV-2 spike protein 1 (S1) IgG is used (Euroimmun, Lübeck, Germany – CE certified version: sensitivity of 91,7% in samples taken 10-20 days after symptoms onset, specificity 88,2-92,4% according to manufacturer). A similar ELISA detecting anti-SARS-COV-2 S1 immunoglobulin A (IgA) is performed in a subset of samples.

To improve sensitivity and specificity an anti-SARS-CoV-2 nucleocapsid protein (NCP) IgG ELISA based on a modified NCP (Euroimmun, Lübeck, Germany – CE certified version: specificity 99.8□%, sensitivity 94.6% after day 10 according to manufacturer) will be employed. For rapid testing, chromatographic lateral flow device in cassette format is employed (‘WANTAI’, SZABO SCANDIC, Vienna, Austria – CE certified version, data sheet (Feb 26th, 2020) reports sensitivity of 95.6% and specificity of 95.2% (199/209)).

### Testing of virus neutralisation

In collaboration with partner laboratories from the German Center for Infection Research, we use an *in vitro* cell culture-based virus neutralisation assay employing plaque assays and clinical SARS-CoV-2 strains from the current pandemic [20]. We also use a novel surrogate virus neutralisation test (sVNT) developed in a partner laboratory detecting the ability of sera to inhibit binding of the receptor-binding domain (RBD) of SARS-CoV-2 to surface-immobilized Angiotensin converting enzyme 2 (ACE2) [21].

### Biobanking and further planned immunological assays

At designated time points, EDTA blood is drawn and PBMCs are isolated and stored in liquid nitrogen for analysis of cellular immune responses to SARS-CoV-2.

Stored samples will be used to study the frequency, phenotype and function of anti-SARS-CoV-2-specific T and B lymphocytes in peptide-based cell stimulation assays. Particularly in participants with confirmed seroconversion, additional tests addressing other aspects of cellular and innate immunity will be conducted.

All blood samples are available for additional research questions, once alternative serology testing systems or experimental tools evaluating functional viral immunity are well-established.

### Possible confounders and bias

Possible confounders or effect modifiers are constantly evaluated during the course of our investigation, and our flexible study design with regular updates of questionnaires and laboratory techniques aims at correcting for these factors. For example, seasonal changes in infection prevalence will influence the rate of airway symptoms possibly related to COVID-19. To correct for this, epidemiological data from public health sources will be taken into account. Another possible confounder is the occurrence of other corona viruses with potential cross reactivity in our serological assays. We plan to correct for this by adapting our testing strategy to assure SARS-CoV-2 specificity. Also the potential emerge of a vaccine could significantly influence our study design, and we aim to correct for this with appropriate questionnaire-based information collection and, if necessary, adapted serological testing. Factors influencing the self-perceived and actual risk of contracting SARS-CoV-2 as HCP such as regional dynamic of COVID-19 cases, season and availability of PPE influence the study results and are as such not regarded as confounders but included into our data collection. To optimize statistical correction of possible confounders, statistical guidance will be sought.

### Study registration, approval by ethical authorities, and informed consent

The study is registered at the German Clinical Trial Register (DRKS00021152) and approved by local authorities. Ethical approval from the Institutional Review Board of Hannover Medical School and data security management approval has been secured (approval # 8973_BO_K_2020). Written informed consent is obtained from all study participants. Each participant receives written information on study procedures and data management. Participants are informed about specimen and data collection, as well as storage of samples for future research projects. Study participation is voluntary and participants have the right to withdraw consent at any time and without disclosure of reasons for withdrawal. Furthermore, they receive a written data security protocol. A trained member of the study team is available for questions at enrolment or at any later time point of participation.

### Protection of personal data

Participant confidentiality is maintained throughout the investigation. All participants receive a pseudonymised study identification number by the investigation team for the labelling of questionnaires and specimens. The link between this identification number and individuals is guarded by a data security trustee and is not disclosed elsewhere. All pseudonyms are replaced by a secondary pseudonym by the study team before data is shared. A data protection protocol has been implemented and approved by the data protection officer at Hannover Medical School. Each participant receives a copy of the data security protocol at enrolment.

### Study size calculation and statistical analysis

Descriptive statistics are calculated employing SPSS^®^ Statistics (Version 26) and GraphPad Prism^®^ (Version 5) and presented as mean +SEM or median/range. Depending on data structure, statistical tests such as Pearson correlation or Fisher’s exact test will be performed and differences between groups will be assessed by t- or ANOVA testing with appropriate post hoc corrections. Statistical guidance will be sought for more complex statistical evaluations and corrections for possible confounders and missing data and losses to follow-up. For the pilot cohort (‘CoCo 1.0’), no formal sample size calculation was performed. In this pilot cohort, an initial prevalence of SARS-CoV-2 IgG antibodies of 0.92% was observed (April 2020). Sample size calculation was performed based on a secondary comparative objective in regard to the regional COVID-19 case number. The known infection rate in the Hannover region at this time point was 0.2%. Assuming that a further 0.2% of the population was infected without detection, the sample size calculation yielded a sample size of n=1,540 (incidence in HCP: 0.92%, incidence in general population: 0.4%, alpha: 0.05, beta 0.2, power 0.8). We therefore set a recruitment goal of at least n=1,550 HCP for the ongoing, prospective study enrolment.

### Reporting of findings

This study will report on serological data and COVID-19 related symptoms, risk factors, and risk-perception in relation to factors such as time, age, sex, profession and type of work as HCP, family circumstances, and others. We will include a short synopsis of the study design in every manuscript and reference this protocol. Further reports will include findings on the magnitude and kinetics of humoral immune responses in HCP against SARS-CoV-2 as well as findings obtained through analysis of questionnaires or biomaterials.

### Communication and participant information

We have set up a study website (www.cocostudie.de) with general information on the study. After personal login, individual study results are available to each participant. For each pseudonym, individual results of the serological tests as well as information on how to interpret the results are communicated to the participants. After evaluation of results of the pilot cohort, every positive or borderline positive anti-SARS-CoV-2 S1 IgG result is confirmed in a second alternative serological testing system prior to providing the validated test result to the participant.

### Funding

This study received seed funding by local patient organizations and clinical and research communities such as the ‘Rheumazentrum’ and ‘Verein Kinderherz Hannover’. It is also supported by grants from industry partners such as Novartis and PARI. The funders have no influence on study design, data collection and analysis, decision to publish, or preparation of manuscripts.

## Discussion

The current SARS-CoV-2 pandemic has challenged healthcare systems and HCP worldwide in a manner unprecedented in modern times [2, 3]. The risk of HCP of contracting the virus appears to strongly depend on the phase of the pandemic, i.e. preparedness of the healthcare system and regional COVID-19 prevalence [3-6].

The CoCo study aims to assess immune responses against SARS-CoV-2 and was initiated in the spring of 2020 as a rapid response to the onset of the pandemic spread in Europe. It was designed as a comparably simple and straightforward observational study that would be adaptable to the rapidly changing dynamics of the pandemic. In the study design, we also focused on feasibility, ensuring that the study would not interfere with provision of routine clinical care and would not increase the risk of HCP to contract SARS-CoV-2. Especially during the initial phase of study setup, resources were scarce and materials such as disinfectant for blood withdrawal and laboratory tubes had to be carefully allocated in order to preserve valuable and potentially limited resources for clinical care. All materials were stocked for six month and need was recalculated for every adaptation of the study schedule. We focused on providing a quick and uncomplicated study experience for participants to avoid disruption of workflows within departments during the pandemic. This initial planning required communication, teamwork, and careful calculation. To test the feasibility of our strategic study design, recruitment for the pilot cohort started immediately in March 2020 [17]. All procedures and protocols were planned for quick evaluation and adaptation with potential to quickly scale up recruitment and testing.

A particular focus in the study design was put on communication with participating HCP. This study was setup to assess the true risk and subjective risk perception of contracting SARS-CoV-2 in clinical staff, and the active involvement of HCP providing feedback on our study design helped us to identify the most relevant questions in this population and to adapt our strategy accordingly. This adaptation included, for example, inclusion of new questions into the questionnaire such as those addressing psycho-social factors and risk perception, as well as the introduction of a second alternative serological test in each case of positive anti-SARS-CoV-2 S1 IgG result.

The study was started during a time when media coverage on the massive overflow of patients and infections in HCP in Italy was pervasive. Germany and its healthcare system was fortunately spared from being overwhelmed as infection rates remained comparably low. We adapted to this situation by stretching the intervals of serological testing in our HCP from weekly screening at the beginning of our pilot cohort enrolment to testing every six months in current enrolment. Given the continuing low seroprevalence amongst HCP in Germany [22, 23], we also actively recruited a control cohort that enabled us to validate our serological assays. During the course of our initial observation period, a plethora of manuscripts describing the immune response against SARS-CoV-2 were published, showing that protective immunity against this virus is a particularly complex immunological process involving multiple immunological factors [15, 24]. Obviously, protective immunity against SARS-CoV-2 cannot be ascertained by serological testing analyses alone, but should be complemented by virus inhibition assays, and, ideally, the assessment of further adaptive and innate cellular and humoral factors. We addressed this by seeking collaboration with researchers on our campus and beyond. During the course of our study, we will continuously strive to identify, record and correct for possible confounders and seek guidance for optimal statistical correction for these factors.

Our interdisciplinary team consisting of specialists in clinical and translational immunology, paediatrics, and infection and respiratory medicine, was instrumental for the swift initiation and continuous development of the study. The quick setup was furthermore supported by the fact that core team of researchers in this study had experience in collaborating with each other during previous local and national healthcare crises such as the influx of refugees in 2015 [25-29]. The rapid development and start the CoCo study, including obtaining of all ethical and legal requirements, acquisition of funding, and all necessary laboratory materials was only possible through extensive teamwork. Initially, the study team was split into groups without physical contact that could substitute for each other in case of an infection or quarantine within the team. To tackle additional challenges and optimize workflows, life science university students were recruited to work on the project.

Given that we are currently experiencing an increase in COVID-19 diagnoses and a decline of the mean age of individuals with COVID-19 [9], our long-term and longitudinal study design is a valuable investment in establishing platforms to explore serological test systems and detecting clinically silent anti-SARS-CoV-2 seroconversions. This ongoing prospective study will answer central questions on prevalence, kinetics, and magnitude of anti-SARS-CoV-2-humoral immune responses and the validity of serological testing of HCP in a region with high healthcare standards and comparatively low COVID-19 prevalence. As such, our results could be highly relevant to other regions and may support HCP around the world in managing this unprecedented situation.

## Supporting information

STROBE checklist

## Data Availability

Technical appendix, statistical code, and dataset available from the corresponding author

## Declarations

### Ethics approval and consent to participate

The study is registered at the German Clinical Trial Register (DRKS00021152) and approved by local authorities. Ethical approval from the Institutional Review Board of Hannover Medical School and data security management approval has been gained and is registered at # 8973_BO_K_2020. Informed consent is obtained from all participants of the study before enrolment into the study. Each participant receives written information on study procedures and data management and the data security concept. The participants are informed about specimen and data collection, as well as storage of samples for future research projects and are informed about their participation being voluntary and their rights to withdraw consent at any time.

### Consent for publication

The manuscript contains no individual person’s data in any form.

### Availability of data and material

The dataset necessary to interpret, replicate and build upon the findings reported in the article will be made available on reasonable request and can be obtained by contacting the corresponding author.

### Authors’ contributions

Research design: AJ, CH, GMNB

Setup of recruitment and laboratory infrastructure: AC, MVS, AZS, DE

Writing and contributing to writing of the manuscript: All authors contributed to writing, have read and approved the manuscript.

## Acknowledgements

We gratefully acknowledge the contributions of Marion Hitzigrath, Melanie Ignacio, Annkathrin Anton, Till Redeker, Madeleine Rommel, Luis Manthey for technical and logistical support, all members of the CoCo Study team for help with enrolment and blood collection (Martina Toussaint, Marcel Winkelmann, Mark Greer, Marcus Bachmann, Birgit Heinisch, Moritz Kayser, Christian Dopfer, Martin Wetzke). Most importantly, the authors would like to thank all study participants recruited thus far for their participation and for their work in healthcare during these stressful times.

## Abbreviations

COVID-19: coronavirus disease 19
SARS-CoV-2: Severe acute respiratory syndrome coronavirus 2
WHO: world health organization

